# Effectiveness of COVID-19 Vaccines: Eight Months Post Single Dose Vaccination

**DOI:** 10.1101/2021.09.18.21263262

**Authors:** Naif Khalaf Alharbi, Jaffar A. Al-Tawfiq, Suliman Alghnam, Amal Alwehaibe, Abrar Alasmari, Suliman A. Alsagaby, Faizah Alotaibi, Faisal Alsubaie, Majid Alshomrani, Fayssal M. Farahat, Mohammad Bosaeed, Ahmad Alharbi, Omar Aldibasi, Abdullah M. Assiri

**Affiliations:** King Abdullah International Medical Research Center (KAIMRC), Riyadh, Saudi Arabia; King Saud bin Abdulaziz University for Health Science (KSAU-HS), Riyadh, Saudi Arabia; Specialty Internal Medicine and Quality Department, Johns Hopkins Aramco Healthcare, Dhahran, Saudi Arabia; Infectious Disease Division, Department of Medicine, Indiana University School of Medicine, Indianapolis, IN, USA; Infectious Disease Division, Department of Medicine, Johns Hopkins University School of Medicine, Baltimore, MD, USA; Department of Infectious Disease Epidemiology, London School of Hygiene & Tropical Medicine, Keppel Street, London WC1E 7HT, UK; Department of Medical Laboratory Sciences, College of Applied Medical Sciences, Majmaah University, Al Majmaah-11952, Saudi Arabia; Assistant Agency for Preventive Health, Ministry of Health, Riyadh, Saudi Arabia; King Abdulaziz Medical City (KAMC), Ministry of National Guard – Health Affairs (MNG-HA), Riyadh, Saudi Arabia

**Keywords:** Effectiveness, COVID-19, vaccines, Pfizer-BioNtech BNT162b2 and AstraZeneca-Oxford AZD1222

## Abstract

**Objectives:** To describe the real-world data on the effectiveness of Pfizer-BioNtech BNT162b2 and AstraZeneca-Oxford AZD1222 vaccines against COVID-19 in a large cohort in the Kingdom of Saudi Arabia (KSA).

**Methods:** A total of 18,543 subjects received a single-dose of either of the vaccines at one vaccination centre in KSA, and were followed up for three to eight months. Clinical data from medical records, adverse events (AEs) from a self-reporting system, and COVID-19 infection data from the national databases were retrieved and analysed.

**Results:** Subjects median age was 33 years old with an average of 27.3 body mass index and the majority were male (60.1%). 92.17% of the subjects had no COVID-19 infection post-vaccination. Diabetes mellitus (p=0.0325), organ transplantation (p=0.0254), and morbid obesity (p=0.0014) were risk factors for infection post-vaccination. Unlike vaccine type, being Saudi, male, or obese was more likely to get the infection earlier. AE reports from 1084 subjects included injection site pain, fatigue, fever, myalgia, headache.

**Conclusion:** Single-dose COVID-19 vaccines in KSA showed an effectiveness rate of 92.17% up to eight months follow-up. The rate for AZD1222 was higher than what have been previously reported. Side effects and AEs were within what has been reported in clinical trials.

## Introduction

A novel Severe Acute Respiratory Syndrome Coronavirus-2 (SARS-CoV-2) emerged in Wuhan, China in 2019 and spread globally (1). The disease is known as coronavirus infectious disease 2019 (COVID-19) (2). In March 2020, the World Health Organization (WHO), declared COVID-19 as a global pandemic and a public health emergency of international concern (3). SARS-CoV-2 is highly contagious and transmitted through human-to-human contact (3). Most COVID-19 cases (81%) are asymptomatic or present with mild to moderate symptoms(4). However, other cases are severe (14%) to critical pneumonia (5%); with a fatality rate of 2-3% (4). As of 5^th^ of September 2021, more than 222 million confirmed cases of COVID-19 and 4.5 million deaths were reported globally (5). To reduce the risk of SARS-CoV-2 transmission, preventive strategies have been implemented, including the use of face masks, hand washing/hygiene, contact tracing, travel bans, and government-led cancellation of unnecessary activities (6).

Importantly, prophylactic vaccines are sought as the ultimate intervention to bring the pandemic under control. Over 200 vaccine candidates for COVID-19 were at various stages of clinical development (7). Of these, at least 50 candidate vaccines made it into human trials. To date, several vaccines have been approved by regulatory authorities, based on clinical trials that demonstrated a high safety profile and variable efficacy rates (7). The results of phase III clinical trials showed 95% efficacy for the Pfizer-BioNtech BNT162b2 vaccine, 92% for the Gamaleya SputnikV vaccine, 94.5% for Moderna mRNA-1273 vaccine, 70% for the AstraZeneca-Oxford AZD1222 vaccine, and 97% for Sinopharm BIBP COVID-19 vaccine (7–9).

Several different vaccine technologies have been used to develop the COVID-19 vaccines (10), including mRNA (11,12), adenoviral vectored vaccines (13,14), inactivated virus (15), and adjuvanted spike glycoprotein (16). The BNT162b2 vaccine is based on the lipid nanoparticle– encapsulated nucleoside-modified messenger RNA (mRNA) that encodes the SARS-CoV-2 spike (S) glycoprotein (17), while the AZD1222 vaccine is based on the ChAdOx1 adenoviral vector encoding the SARS-CoV-2 spike (S) glycoprotein (10). The Kingdom of Saudi Arabia (KSA) was among the first countries to launch an accelerated program for COVID-19 vaccination (18). The approved COVID-19 vaccines that are being used in KSA are the Pfizer-BioNtech BNT162b2 vaccine, Moderna mRNA-1273 vaccine, and AstraZeneca-Oxford AZD1222 vaccine (19).

For each vaccine candidate, there were a number of vaccine controlled randomised clinical trials focusing on evaluating safety, immunogenicity, and efficacy (7,20), which were the basis for regulatory approvals. However, to gain a better understanding of how COVID-19 vaccines would perform in various populations, it is essential to gather real-world data and analysis post-vaccination, in particular concerning different ethnic populations, in order to establish the long-term effectiveness (protection rate) in several different sub-populations. These subpopulations can be defined based on differences in gender, age, nationality, occupation, comorbidities and chronic disease (including haemodialysis and oncology patients).

Therefore, this study was conducted to evaluate the long-term effectiveness (protection rate) of Pfizer-BioNtech BNT162b2 and AstraZeneca-Oxford AZD1222 COVID-19 vaccines in KSA, including national and expatriate subjects from different countries. It also looks at the demographics and clinical characteristics of subjects and the adverse events (AE) post-vaccination.

## Methods

### Subjects and data collection

In this study, data for 20555 vaccinated subjects were collected from a single vaccination centre at National Guard Health Affairs (NGHA) hospitals in Riyadh city, Saudi Arabia. The subjects received either BNT162b2 or AZD1222 vaccines. Clinical data for all subjects were retrieved from the MNG-HA medical records. Confirmation of COVID-19 infections were obtained from the national database at the Ministry of Health which covers any COVID-19 PCR testing performed in the country. Subjects were given access to an online portal for reporting symptoms and adverse events; in addition, vaccine safety records at the Infection Prevention and Control Department at the King Abdulaziz Medical City (KAMC), MNG-HA, were interrogated. Side effects reports were obtained for around 6% of the vaccinated subjects. Those who were infected prior to vaccination, had two doses of Pfizer vaccine (who were less than 100 subjects), had the COVID-19 infection within two weeks of vaccination, or lack data on COVID-19 testing were excluded from the analysis presented in this article. Therefore, 18543 out of 20555 subjects were included in the study analysis.

### Statistical Analysis

Descriptive statistics were applied to summarise the data of subjects’ demographical and clinical characteristics and the number of adverse events. Comparison between the two study groups (No infection post-vaccination Infection post-vaccination) in terms of demographical and clinical variables, including gender, nationality, and comorbidities where tested using chi-square. Multivariate analysis was applied to model the probability of infection after vaccination using backward elimination logistic regression with alpha level 0.5 to enter the model and 0.05 to stay in the model for all demographical and clinical variables. The final model is reported in terms of odds ratios and 95% confidence intervals. Using the Wilcoxon rank-sum test, we test the number of days between vaccination and infection for infected individuals in terms of gender, nationality, and comorbidities. All statistical analyses were performed using SAS 9.4 (SAS Institute Inc., Cary, NC) and data visualisation using GraphPad Prism V8 software (GraphPad Software, San Diego, CA).

### Ethical approval

The study was approved by the IRB at KAIMRC for projects RC20/180 and NRC21R-120-03.

## Results

### Clinical and characteristics of the study cohort

Data for a total of 18543 subjects were collected from the medical records at King Abdulaziz Medical City, MNG-HA, Riyadh city. Due to vaccine supply issues as well as to achieve a quick rollout of COVID-19 vaccines, KSA has decided to postpone the second dose of COVID-19 vaccines; therefore, all of the study subjects received only a single dose of either BNT162b2 or AZD1222 vaccines between 19^th^ December 2020 and 14^th^ April 2021. Of the included vaccinees, 410 (2.3%) received BNT162b2 and 18133 (97.8%) received AZD1222. Of the total number of vaccinees, 11145 (60.1%) were males and 7398 (39.1%) were females. The median age was 33 years (IQR: 26 - 42), ranging from 18 to 112 years old; and the median body mass index (BMI) was 27.3 (IQR: 23.8 - 31.4). Results of SARS-CoV-2 PCR tests were obtained from the national database at the Ministry of Health up 10^th^ August 2021; meaning that individuals were followed up for at least three months and almost eight months at maximum.

### Effectiveness of COVID-19 vaccines

Vaccinated subjects who have no documented COVID-19 infection prior to vaccination were analysed in order to provide real-world data on COVID-19 vaccine effectiveness in this cohort. Subjects were followed up for at least three months (last vaccination was on the 14^th^ April and last follow up was on the 10^th^ August). Out of 18543, 17091 (92.17%) remained uninfected post-vaccination while the infection was documented for 1452 (7.83%) of the vaccinated individuals, Table 1. Forty-six (11.2% of those who received BNT162b2) and 1406 (7.75% from those who received AZD1222) had the infection post two weeks of the single dose vaccination, indicating the effectiveness of these vaccines, Table S1. Age does not appear to vary significantly in the two groups of vaccinated subjects (uninfected and infected); median age was 33 (IQR: 26 - 43) for the uninfected group and 33 (IQR: 26 - 41) for the infected group. Likewise, the BMI was similar in the uninfected group (median = 27.3 (IQR: 23.8 - 31.3) and the infected group (median = 28.1 (IQR: 24.5 - 32.3). In an attempt to identify risk factors associated with infection post-vaccination, we analysed the comorbidity data of the subjects in the two groups (Table 1). Male gender and Saudi nationality appeared to have more infection post-vaccination than female and non-Saudi nationality (p<0.0001). Diabetes (p=0.0325), organ (mainly kidney) transplantation (p=0.0254), and morbid obesity (p=0.0014) were found to be associated with the risk of infection in vaccinated subjects. Remarkably, Lung diseases, asthma, or cancer, for which treatment by chemotherapy predisposes patients to microbial infection due to leukocytopenia, did not appear as risk factors of infection in vaccinated subjects. A multivariate logistic regression analysis on infection post-vaccination, demographics and co-morbidities showed that Saudi, male, and obese subjects are more likely to contract COVID-19 post a single dose of COVID-19 vaccines (Table 2). Infections that occur within two weeks of vaccination were only 73 cases and were not included in the study analysis. The data showed that the time between vaccination and infection (excluding the 73 cases) was between 14 and 146 days (median = 82 days), Figure 1. These periods of time were not different when broken by vaccine type (data as per vaccine type is not shown) and COVID-19 cases following the BNT162b2 vaccine were only 46, which does not allow a powerful statistical conclusion on this regard. In addition, these times were not different with regard to subject comorbidities (Table S2).

**Table 1:** Co-morbidities and other characteristics of subjects vaccinated with either BNT162b2 or AZD1222 COVID-19 vaccines.

|  | No infection post-vaccination | Infection post-vaccination | P value |
| --- | --- | --- | --- |
|  | (n=17091) | (n=1452) |  |
| **Male** | 10188 (59.62%) | 957 (65.91%) | <0.0001 |
| **Female** | 6903 (40.38%) | 495 (34.09%) |  |
| **Nationality** | Saudi= 12026 (70.36%) | Saudi= 1189 (81.89%) | <0.0001 |
|  | Non-Saudi= 5065 (29.64%) | Non-Saudi= 263 (18.11%) |  |
| **Diabetes mellitus** | 1602 (9.37%) | 161 (11.09%) | 0.0325 |
| **Hypertension** | 1916 (11.21%) | 62 (11.16%) | 0.9505 |
| **Hyperlipidemia** | 1009 (5.90%) | 77 (5.30%) | 0.3494 |
| **Chronic kidney disease** | 213 (1.25%) | 18 (1.24%) | 0.9826 |
| **Chronic lung disease** | 604 (3.53%) | 61 (4.20%) | 0.1894 |
| **Asthma** | 578 (3.38%) | 58 (3.99%) | 0.2182 |
| **Malignancy** | 183 (1.07%) | 16 (1.10%) | 0.9118 |
| **Morbid obesity** | 603 (3.53%) | 75 (5.17%) | 0.0014 |
| **Haemodialysis** | 93 (0.54%) | 8 (0.55%) | 0.973 |
| **Organ Transplant** | 11 (0.06%) | 4 (0.28%) | 0.0254 |

**Table 2:** Odd ratios from a multivariate logistic regression analysis modelling the probability of COVID-19 infection post-vaccination.

| **Variable** | **Odds Ratio** | **CI** | **P Value** |
| --- | --- | --- | --- |
| Gender: Male | 1.167 | 1.039-1.311 | 0.0091 |
| Nationality: Saudi | 1.805 | 1.567-2.079 | <0.0001 |
| Obese | 1.327 | 1.033-1.705 | 0.0271 |

**Figure 1:**
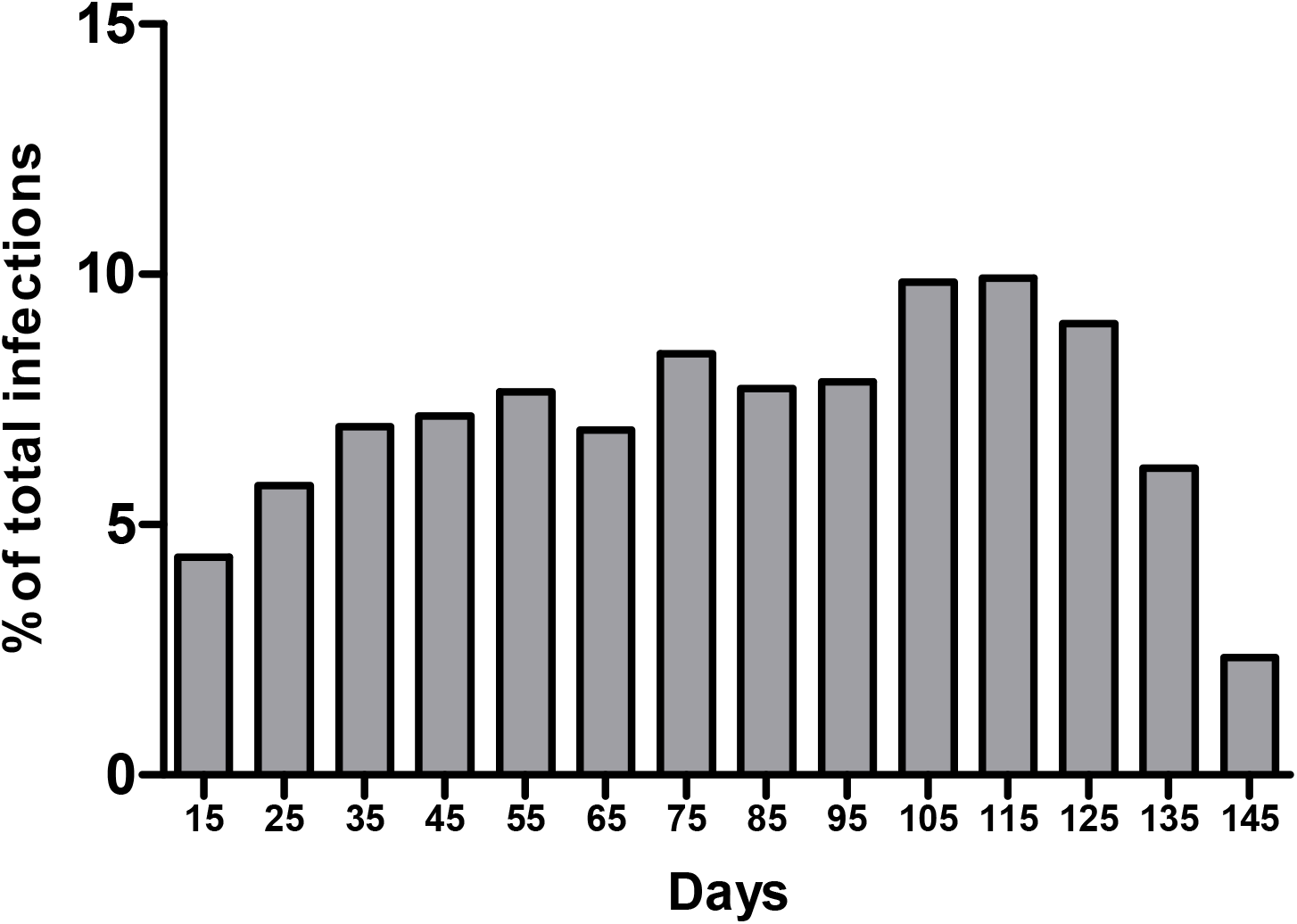
Days between COVID-19 vaccination and infections shown as percentage of the infected subjects.

### Adverse events to COVID-19 vaccines

To provide insights on the adverse events (AE) to the vaccines in the study subjects, only 1084 subjects utilised the online reporting portal or contacted NGHA hospitals to report AE (Table 3). Although around 5.1% of the vaccinated subjects reported AE, lack of reporting does not ensure lack of side effects. Injection site pain was the most frequently reported AE (800 cases). Other common AEs include fatigue (732 cases), fever (714 cases), myalgia (678), headache (657 cases), joint pain and malaise (399 cases). In contrast, AEs, such as skin rash, cough, abdominal pain and tachycardia were only reported by single individuals among vaccinated subject.

**Table 3:**
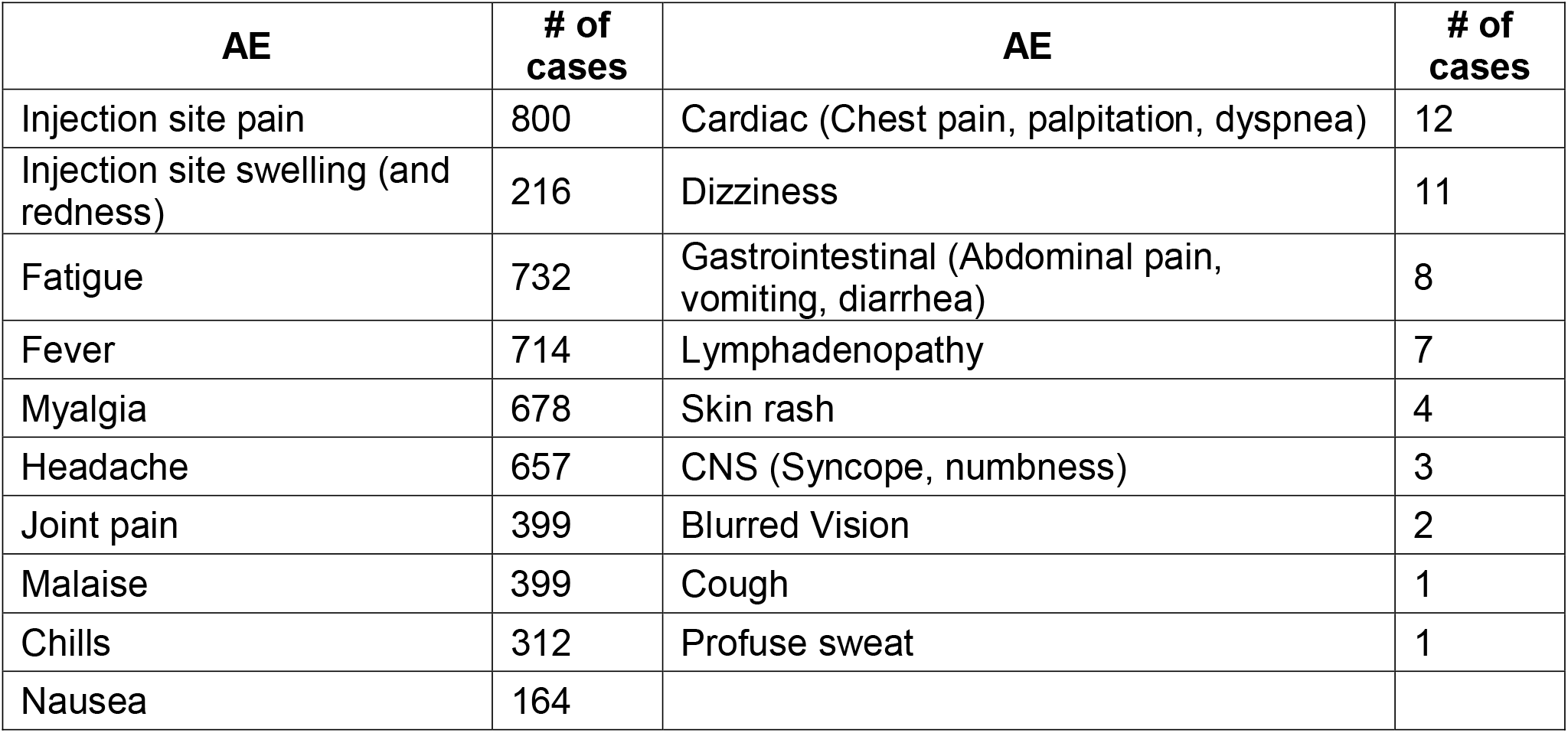
Reported adverse events post COVID-19 vaccination.

| **AE** | **# of cases** | **AE** | **# of cases** |
| --- | --- | --- | --- |
| Injection site pain | 800 | Cardiac (Chest pain, palpitation, dyspnea) | 12 |
| Injection site swelling (and redness) | 216 | Dizziness | 11 |
| Fatigue | 732 | Gastrointestinal (Abdominal pain, vomiting, diarrhea) | 8 |
| Fever | 714 | Lymphadenopathy | 7 |
| Myalgia | 678 | Skin rash | 4 |
| Headache | 657 | CNS (Syncope, numbness) | 3 |
| Joint pain | 399 | Blurred Vision | 2 |
| Malaise | 399 | Cough | 1 |
| Chills | 312 | Profuse sweat | 1 |
| Nausea | 164 |  |  |

## Discussion

In this study, the long-term effectiveness and self-reported adverse events post-vaccination with two approved COVID-19 vaccines in KSA were evaluated. These vaccines were the Pfizer-BioNtech BNT162b2 and AstraZeneca-Oxford AZD1222 COVID-19 vaccines. The effectiveness of the BNT162b2 and AZD1222 COVID-19 vaccines is very important for the control of the disease. The current study showed a high rate of protection against infection (92%) as only 1452 (7.83%) vaccinees had a COVID-19 infection post-vaccination. After excluding any infection that occurred within 2 weeks of single dose vaccinations, the time between vaccination and infection varied from 14 and 146 days (median = 82 days). In a previous study from KSA, the effectiveness of the AZD1222 vaccine was 99.5% within 3 weeks of the first dose (21). The efficacy of BNT162b2 vaccine in preventing symptomatic COVID-19 infection was 52% (95% CI:□30–86%) after one dose and 95% (95% CI: 90–98%) after two doses in clinical trial settings (8). On the other hand, clinical trials showed an efficacy of 70% (95% CI:□55–81%) after two doses of the AZD1222 vaccine (8). Data from real-world vaccination showed that two doses of BNT162b2 vaccine reduced the risk of SARS-CoV-2 infection by 90% (22). In addition, it was found that most breakthrough infections were either asymptomatic or mildly symptomatic (23).

In the current study, infection post vaccination were significantly higher in subjects with diabetes mellites, obesity, or organ transplantation. In a previous study, the rate of breakthrough infections was not different among vaccinees by age group, gender, or type of vaccine but were lower among those without comorbidities (0.44 [95% CI 0.25,0.62]) compared with those with 1 to 3 comorbidities (24). The contribution of underlying disease to breakthrough infections is of particular importance as additional boosting doses might be required. In one study, the immune responses after COVID-19 vaccine was lower among males and those with underlying comorbidities such as diabetes mellitus, hypertension and renal disease (25). A meta-analysis showed that diabetic patients had increased risk for severe COVID-19 and increased mortality (26). A multivariate logistic regression analysis on infection post-vaccination in the current study showed that Saudi, male, and obese subjects were more likely to contract COVID-19 post a single dose of COVID-19 vaccines. The reason for Saudi male to have higher rate of breakthrough infections might be related to the social norms, occupations, and behavioural interactions, warranting further social-epidemiological studies. The contribution of different ethnicity to the development of breakthrough infection was also reported previously (24,27,28).

The percentage of the infected patients in our study would be affected by the endemicity of the virus in the study areas. Additionally, the pandemic has shown frequent waves and variations in the numbers of the daily new cases of COVID-19, which would certainly influence our findings. Emerging variants of the SARS-CoV-2 might have an impact on the vaccine effectiveness, especially for the one dose protection, a strategy that has been followed by several countries due to vaccine logistic issues and the need to expand vaccination coverage in populations. Therefore, long-term follow-up on the vaccinees is needed to estimate the longevity of the single-dose vaccine effect and effectiveness.

Of the 20555 vaccinated subjects, 1084 (5.1%) had utilised the online portal or contacted the hospital to report AEs following the vaccination. In an earlier study on vaccine safety and reactogenicity from KSA, 34.7% reported an adverse event on the first call after the vaccination (21). These differences in the rate of AEs are likely related to the methodology of the two studies. The current study utilised self-reporting and the previous study utilised an active methodology by calling subjects to record any AEs. In the current study, the most reported Aes were injection site pain (n= 800), fatigue (n=732), fever (n=714), myalgia (n=678) and headache (n=657). The previously cited study from KSA also found that local AEs at the injection site were most common after AZD1222 (30.5%) (21) while the current study cannot establish a rate for this AE due to the used method. In a phase III clinical trial, injection-site pain was reported by 48.6% (29); however, this rate of occurrence varied for different vaccines (30).

Fever was the most frequently reported adverse effects after COVID-19 vaccination in this study. Two previous studies from KSA reported fever in 18.5% after the first dose of BNT162b2 vaccine, 1.3% after the second dose and an overall rate of 4.3% (31). The second study reported a rate of 31% after the AZD1222 vaccine (21). In a third study from KSA, fever was reported in 66% of the subjects who received either AZD1222 or BNT162b2 vaccine (32) while fever was reported in 22% of participants receiving BNT162b2 vaccine in a randomised-controlled trial (33). The current study showed that 7 vaccinees had lymphadenopathy after COVID-19 vaccination. The incidence of ipsilateral axillary reactive lymphadenopathy following mRNA vaccine was 13% among 68 patients who had CT-scan (34). In addition, 20 vaccinees developed ipsilateral supraclavicular lymphadenopathy in a separate study (35). The estimated occurrence of lymphadenopathy was 1% and 10% after the after BNT162b2 and mRNA-1273 vaccines, respectively (36,37). Nevertheless, lymphadenopathy is usually documented following other intramuscular vaccines such as influenza and human papillomavirus vaccines (38,39).

In conclusion, this study is the first to investigate the safety and efficacy of two main vaccines in a large cohort in Saudi Arabia including national and expatriate subjects from different countries. Single-dose COVID-19 vaccines showed an effectiveness rate of 92% with no major side effects during the three to eight months observational period.

## Data Availability

Analysed data are presented in this article, but raw data are available upon request.

## Acknowledgments

The authors are grateful for the assistance of the COVID-19 vaccination center at MNG-HA and the Saudi Ministry of Health.

## Supplementary tables

**Table S1:** Co-morbidities and other characteristics of the study subjects (as in Table 1) presented according to the vaccine type.

|  | Vaccination with Pfizer BNT162b2 COVID-19 Vaccine |  |  | Vaccination with AstraZeneca AZD1222 COVID-19 Vaccine |  |  |
| --- | --- | --- | --- | --- | --- | --- |
|  | No infection post-vaccination | Infection post-vaccination | P value | No infection post-vaccination | Infection post-vaccination | P value |
|  | (n=364) | (n=46) |  | (n=16727) | (n=1406) |  |
| **Male** | 213 (63.96%) | 31 (70.45%) | <0.0001 | 9953 (59.54%) | 924 (65.77%) | <0.0001 |
| **Female** | 120 (36.04%) | 13 (29.55%) |  | 6763 (40.46%) | 481 (34.23%) |  |
| **Nationality** | Saudi= 318 (87.4%) | Saudi= 43 (93.5%) | 0.2283 | Saudi= 11708 (69.99%) | Saudi= 1146 (81.51%) | <0.0001 |
|  | Non-Saudi= 46 (12.6%) | Non-Saudi= 3 (6.5%) |  | Non-Saudi= 5019 (30.01%) | Non-Saudi= 260 (18.49%) |  |
| **Diabetes** | 60 (16.5%) | 2 (4.3%) | 0.0304 | 1542 (9.22%) | 159 (11.31%) | 0.0098 |
| **Hypertension** | 56 (15.4%) | 4 (8.7%) | 0.2265 | 860 (11.12%) | 158 (11.24%) | 0.8927 |
| **Hyperlipidemia** | 20 (5.5%) | 1 (2.2%) | 0.492 | 989 (5.91%) | 76 (5.41%) | 0.4372 |
| **Renal diseases** | 35 (9.6%) | 2 (4.3%) | 0.4089 | 178 (1.06%) | 16 (1.14%) | 0.7961 |
| **Lung diseases** | 22 (6.0%) | 1 (2.2%) | 0.4947 | 582 (3.48%) | 60 (4.27%) | 0.1246 |
| **Asthma** | 21 (5.8%) | 1 (2.2%) | 0.492 | 557 (3.33%) | 57 (4.05%) | 0.1494 |
| **Cancer** | 6 (1.6%) |  | 1 | 177 (1.06%) | 16 (1.14%) | 0.7794 |
| **Morbid obesity** | 18 (4.9%) | 6 (14.0%) | 0.0402 | 585 (3.50%) | 69 (4.91%) | 0.0065 |
| **Haemodialysis** | 32 (8.8) | 2 (2.2) | 0.4042 | 61 (0.36%) | 6 (0.43%) | 0.7126 |
| **Transplant** | 1 (0.3%) | 0 (0%) |  | 10 (0.06%) | 4 (0.28%) | 0.0192 |

**Table S2:**
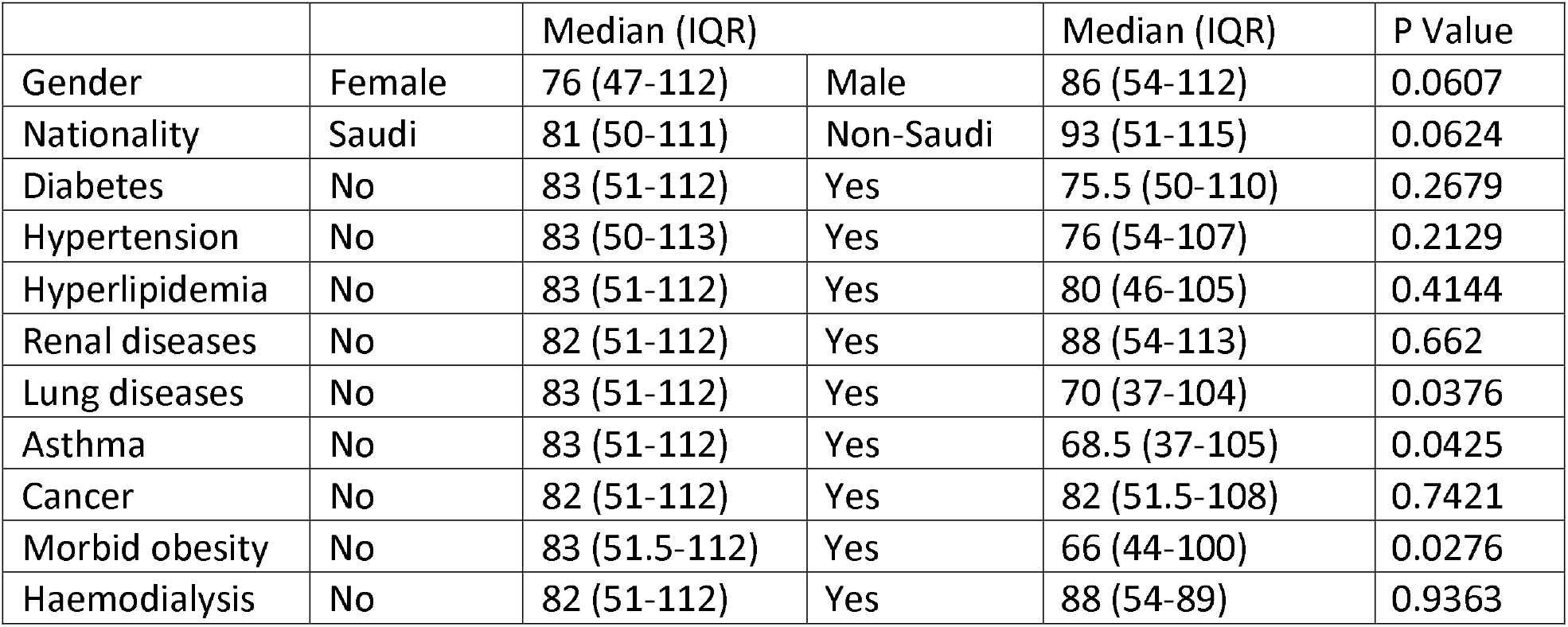
Number of days between vaccination and infection measured for gender, nationality, and comorbidities. Median (and IQR) of the number of days between vaccination and infection analysed according to co-morbidities and other characteristics of the subjects who contracted COVID-19 post-vaccination; excluding infections within two weeks of vaccination.

